# The AFRIDIARRHEA multimodal fusion framework for Estimating the Burden of Diarrheal Diseases Among Children Under Five in Kenya, Zimbabwe, and Somaliland

**DOI:** 10.64898/2026.06.01.26354632

**Authors:** John Onyango Agumba, Lucy Namusonge, Joshua Ogendo, Musiiwa Takavarasha, Mohamad Ahmed Hassan, Morris Senghor, Lydia Waswa

## Abstract

**Background:** Accurate estimation of childhood diarrheal disease burden in Africa remains challenging because of limited surveillance, incomplete mortality data, pathogen-attribution uncertainty, and complex environmental and socioeconomic drivers. This study developed the African Diarrheal Disease Integrated Risk Intelligence and Burden Estimation Architecture (AFRIDIARRHEA), a multimodal fusion framework for estimating under-five diarrheal burden in resource-constrained settings.

**Methods:** AFRIDIARRHEA integrates Bayesian epidemiological modeling, machine learning, temporal forecasting, geospatial analytics, pathogen attribution, environmental intelligence, and uncertainty quantification within a unified framework. Synthetic datasets representing Kenya, Zimbabwe, and Somaliland were used to evaluate mortality, morbidity, hospitalization burden, pathogen-attributed mortality, and predictive performance.

**Results:** The framework identified substantial heterogeneity in disease burden across countries, with Zimbabwe exhibiting the highest modeled mortality and morbidity burden and Somaliland the highest hospitalization burden. Rotavirus and Shigella were the dominant contributors to pathogen-attributed mortality. The multimodal fusion model outperformed the Bayesian baseline and individual component models, achieving improved predictive accuracy, robust uncertainty calibration, and strong agreement with benchmark estimates.

**Conclusions:** AFRIDIARRHEA demonstrates the potential of multimodal fusion modeling for integrated estimation of childhood diarrheal burden, pathogen attribution, and uncertainty in African settings. The framework provides a scalable, transparent, and policy-relevant approach for supporting vaccine prioritization, WASH investments, outbreak preparedness, and child survival programs in data-limited environments.

## 1. Introduction

Diarrheal diseases remain among the leading causes of morbidity and mortality among children under five years of age despite substantial improvements in vaccination coverage, nutrition, water and sanitation infrastructure, and access to oral rehydration therapy. According to the World Health Organization [1], diarrheal diseases account for approximately 443,000 deaths annually among children under five worldwide, with the overwhelming majority occurring in low- and middle-income countries (LMICs), particularly in sub-Saharan Africa and South Asia. Beyond mortality, diarrheal diseases contribute substantially to childhood morbidity, hospitalizations, impaired growth, malnutrition, cognitive deficits, and disability-adjusted life years (DALYs), imposing a significant burden on healthcare systems and socioeconomic development [2]. Although global diarrheal mortality has declined substantially since 1990, the burden remains unevenly distributed across regions and populations. Recent Global Burden of Disease (GBD) analyses indicate that sub-Saharan Africa continues to experience disproportionately high rates of diarrheal morbidity and mortality due to persistent challenges related to unsafe water, inadequate sanitation, undernutrition, limited healthcare access, and climate vulnerability [2]. UNICEF estimates that diarrheal diseases remain responsible for approximately 9% of all deaths among children under five globally, underscoring the continuing need for improved surveillance, prevention, and intervention strategies [3]. Accurate estimation of diarrheal disease burden remains a major epidemiological challenge. Existing burden estimates often differ substantially across organizations and modeling frameworks due to variations in mortality envelopes, etiologic attribution methods, assumptions regarding pathogen prevalence, handling of coinfections, surveillance coverage, and geographic resolution. These challenges are particularly pronounced in LMICs, where weak Civil Registration and Vital Statistics (CRVS) systems contribute to underreporting, incomplete mortality attribution, and limited availability of high-quality epidemiological data [4]. Furthermore, many burden estimation frameworks rely heavily on fragmented clinical surveillance datasets rather than comprehensive mortality data, thereby introducing additional uncertainty into disease burden estimates.

A critical component of diarrheal burden estimation is etiologic attribution, which seeks to determine the proportion of disease cases, hospitalizations, and deaths attributable to specific pathogens. Pathogen-specific burden estimation remains difficult because diarrheal diseases are caused by diverse bacterial, viral, and parasitic pathogens whose prevalence varies across geographic locations, seasons, and population groups. Imperfect diagnostics, asymptomatic carriage, multiple pathogen detections, sparse mortality data, and heterogeneity in laboratory methodologies further complicate attribution efforts [5]. Consequently, deterministic attribution approaches may inadequately represent the complex relationships between pathogen detection and clinical disease outcomes. Major epidemiological investigations have substantially advanced understanding of pathogen-specific diarrheal disease burden. The Global Enteric Multicenter Study (GEMS) and the Malnutrition and Enteric Disease (MAL-ED) study demonstrated that multiple enteric pathogens frequently coexist within diarrheal samples and that probabilistic attribution approaches are often necessary to estimate pathogen-specific contributions accurately [6, 7]. These studies consistently identified Rotavirus, *Shigella, Cryptosporidium*, enterotoxigenic *Escherichia coli* (ETEC), and Norovirus as major contributors to severe childhood diarrheal disease in LMICs. More recent analyses have confirmed that Rotavirus remains one of the leading causes of under-five diarrheal mortality globally despite substantial expansion of vaccine coverage [8]. Cholera continues to contribute significantly to disease burden in endemic regions and during outbreaks, particularly in settings characterized by inadequate water and sanitation infrastructure.

In addition to pathogen-specific factors, environmental and climatic conditions play a critical role in shaping diarrheal disease transmission dynamics. Rainfall variability, flooding, drought, temperature fluctuations, water contamination, and sanitation conditions influence pathogen survival, environmental persistence, and transmission pathways [9]. Numerous studies have demonstrated strong associations between extreme weather events and increased diarrheal disease incidence, highlighting the importance of incorporating environmental intelligence into disease burden estimation frameworks. Recent advances in remote sensing technologies have enabled the use of satellite-derived environmental indicators, including rainfall estimates, vegetation indices, flood extent, land-surface temperature, and surface-water distribution, to support infectious disease forecasting and environmental risk assessment. However, integration of these environmental data streams into diarrheal burden estimation remains relatively limited. Spatial heterogeneity further complicates disease burden estimation across Africa. Diarrheal disease hotspots frequently coincide with regions characterized by poor sanitation, informal settlements, limited healthcare access, climate vulnerability, and high rates of childhood undernutrition. Geospatial epidemiological methods provide important tools for identifying regional heterogeneity and improving geographic targeting of public health interventions. Bayesian spatial models have been widely employed in disease mapping because of their ability to incorporate uncertainty and hierarchical dependencies. Nevertheless, traditional statistical approaches may be limited in their ability to capture highly nonlinear interactions among environmental, demographic, nutritional, and healthcare-related determinants of disease burden.

Recent advances in machine learning have created new opportunities for improving disease burden estimation and forecasting. Machine-learning algorithms such as Extreme Gradient Boosting (XGBoost) and Light Gradient Boosting Machine (LightGBM) have demonstrated strong performance in capturing complex nonlinear relationships among environmental, demographic, and clinical variables [10, 11]. These approaches offer important advantages over conventional linear models when modeling multifactorial disease systems characterized by heterogeneous drivers and interactions. In parallel, multimodal fusion architectures have emerged as powerful frameworks for integrating information from multiple data sources and modeling paradigms. Such architectures can combine mechanistic epidemiological models, Bayesian inference systems, geospatial analytics, temporal forecasting models, environmental-risk indicators, and machine-learning algorithms within a unified analytical framework. Despite growing adoption of multimodal approaches in infectious disease forecasting, relatively few studies have developed comprehensive frameworks specifically designed for African diarrheal disease burden estimation. Existing investigations often focus on isolated aspects of disease modeling, including pathogen attribution, geospatial mapping, climate forecasting, or statistical burden estimation, rather than integrating these complementary approaches into a single decision-support system. Consequently, substantial opportunities remain to improve burden estimation through the integration of epidemiological, environmental, spatial, temporal, and machine-learning information streams.

To address these limitations, the present study developed the African Diarrheal Disease Integrated Risk Intelligence and Burden Estimation Architecture (AFRIDIARRHEA), a multimodal fusion framework designed to estimate diarrheal mortality, morbidity, pathogen-attributed burden, and associated uncertainties among children under five years of age in Kenya, Zimbabwe, and Somaliland. The framework integrates Bayesian epidemiological modeling, machine-learning algorithms, geospatial analytics, temporal forecasting, environmental-risk assessment, and pathogen attribution methodologies within a unified architecture. By combining diverse data streams and analytical approaches, AFRIDIARRHEA seeks to provide policy-relevant, geographically resolved, and uncertainty-aware estimates capable of supporting evidence-based public health planning and intervention prioritization in resource-constrained settings.

## 2. Methods

### 2.1. Study Design

This study implemented a multimodal proof-of-concept burden-estimation framework using synthetic datasets representing Kenya, Zimbabwe, and Somaliland for the year 2025 as shown in Figure 1. The synthetic datasets were designed to emulate realistic epidemiological, environmental, geospatial, and pathogen-specific relationships relevant to under-five diarrheal disease transmission in Kenya, Somaliland and Zimbabwe.

**Figure 1.**
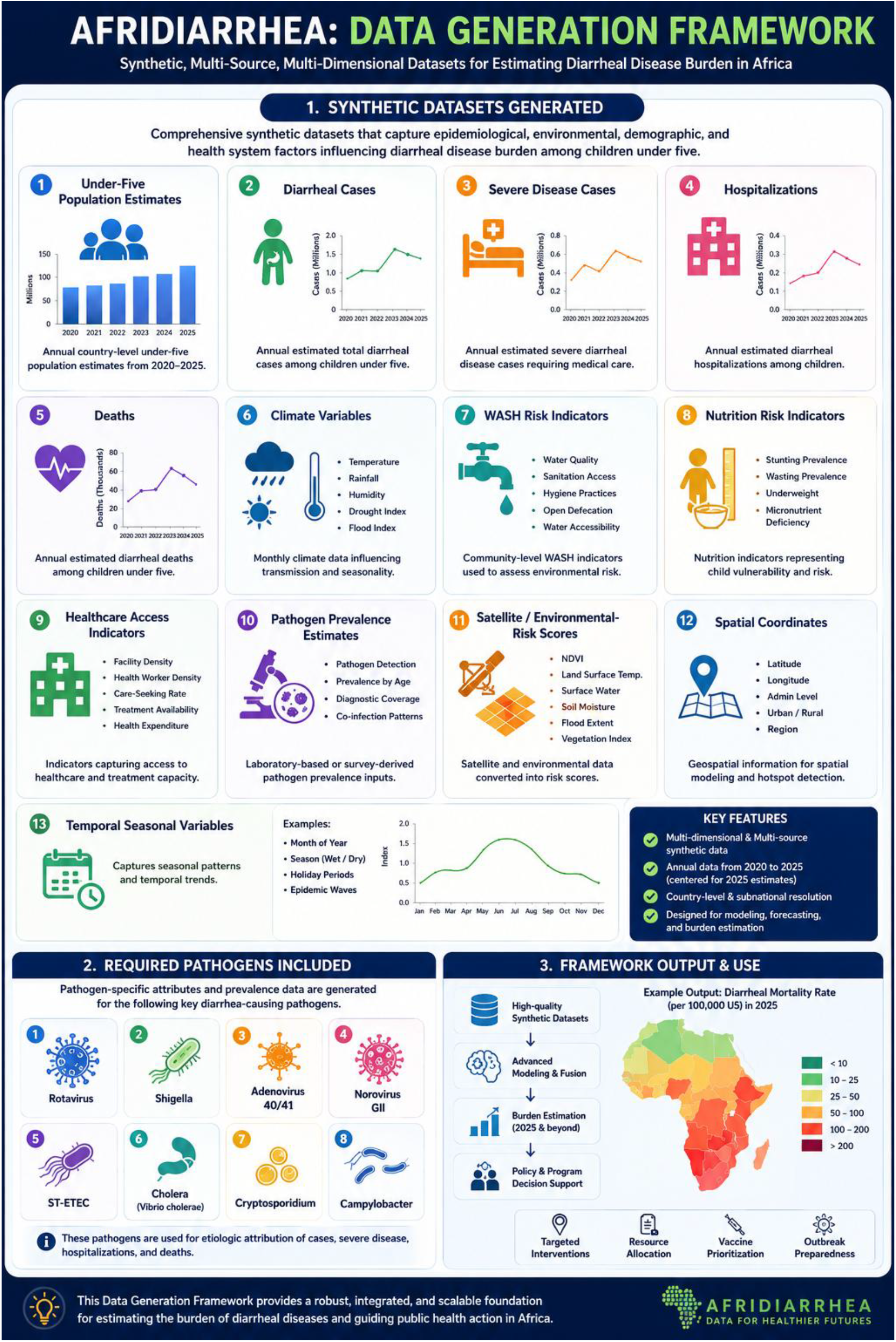
AFRIDIARRHEA data generation framework showing the overview of the synthetic multi-source data generation architecture used to estimate diarrheal disease burden among children under five in Africa, integrating epidemiological, environmental, climatic, healthcare, geospatial, and pathogen-specific variables for burden estimation, forecasting, and policy decision support The framework was developed in Python using epidemiological modeling, machine-learning algorithms, geospatial features, temporal predictors, and uncertainty calibration.

### 2.2. Data Generation Framework

Synthetic datasets included under-five population estimates, diarrheal cases, severe disease cases, hospitalizations, deaths, climate variables, WASH risk indicators, nutrition risk indicators, healthcare access indicators, pathogen prevalence estimates, satellite/environmental-risk scores, spatial coordinates and temporal seasonal variables. Required pathogens included Rotavirus, Shigella, Adenovirus 40/41, Norovirus GII, ST-ETEC, Cholera, Cryptosporidium and Campylobacter.

### 2.3. Bayesian Hierarchical-Style Baseline

The baseline model represented diarrheal burden as Equation (1):

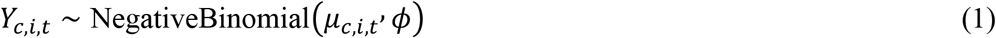

where *Y*_*c,i,t*_ represented observed burden, *μ*_*c,i,t*_ represented expected burden and *ϕ* represented overdispersion

The expected burden was modeled as Equation (2):

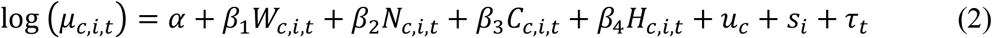

where *W*_*c,i,t*_ is WASH risk, *N*_*c,i,t*_ is nutrition risk, *C*_*c,i,t*_ is climate risk, *H*_*c,i,t*_ is healthcare access, *v*_*c*_ is country-level effect, *s*_*i*_ is spatial effect and *τ*_*t*_ is temporal effect.

### 2.4. Machine-Learning Models

The XGBoost-style model was represented as:

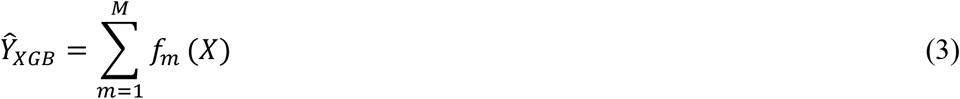

The LightGBM-style model was represented as:

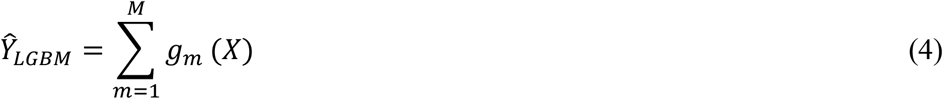

### 2.5. Temporal Forecasting

Seasonality and lag effects were represented as:

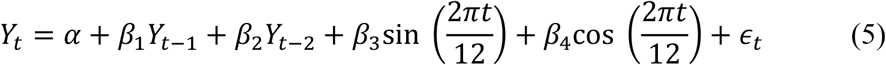

### 2.6 Geospatial Modeling

Spatial risk was represented as:

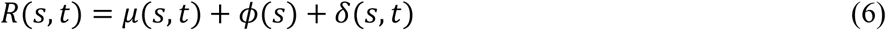

### 2.7. Pathogen Attribution

Attributable fractions were estimated using Equation (7):

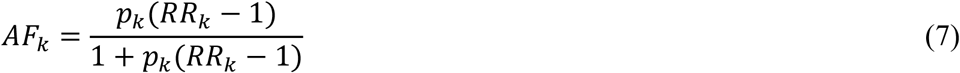

where *AF*_*k*_ represent the attributable fraction for pathogen *k, p*_*k*_ represent the pathogen prevalence and *RR*_*k*_ represent the pathogen relative risk

Pathogen-attributed deaths were estimated as Equation (8):

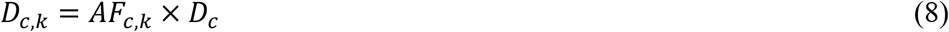

### 2.8. Coinfection Adjustment

Coinfection probability was represented as Equation (9):

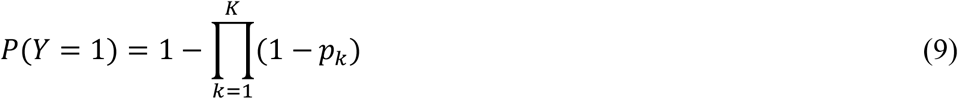

### 2.9. Fusion Model

The multimodal fusion model combined all model families as Equation (10):

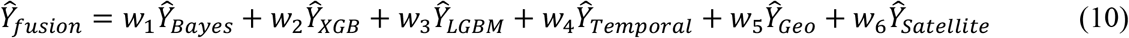

Weights satisfied Equation (11):

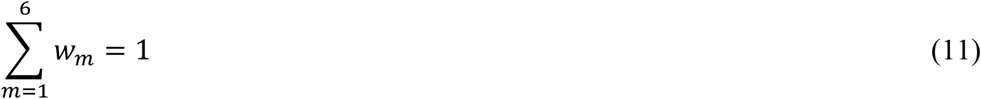

### 2.10. Uncertainty Calibration

The calibrated uncertainty was represented as:

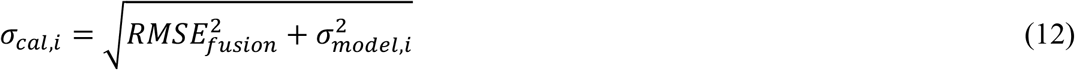

The 95% uncertainty interval was represented as:

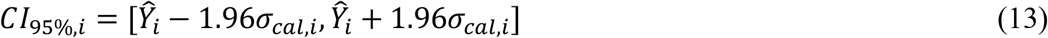

### 2.11. Evaluation Metrics

Model performance was evaluated using Mean Absolute Error (MAE), Root Mean Square Error (RMSE) and coefficient of determination (*R*^2^).

## 3. Results

### 3.1. Country-Level Burden Estimates

Table1 presents the country-level estimates of under-five diarrheal disease burden generated by the AFRIDIARRHEA framework for Kenya, Somaliland, and Zimbabwe. The table summarizes key epidemiological indicators, including under-five population, total diarrheal cases, severe cases, hospitalizations, deaths, mortality incidence rates, and morbidity incidence rates.

**Table 1.**
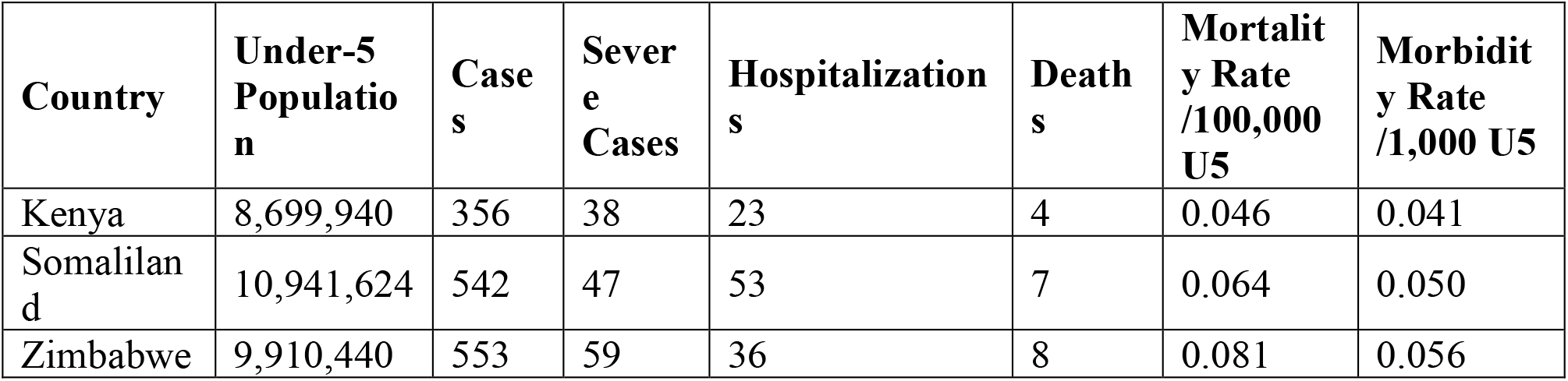
Country-level estimates of under-five diarrheal morbidity, mortality, severe cases, hospitalizations, and incidence rates generated by the AFRIDIARRHEA framework for Kenya, Somaliland, and Zimbabwe in 2025.

The country-specific burden estimates revealed substantial heterogeneity in diarrheal disease outcomes across Kenya, Zimbabwe, and Somaliland. The results show that Zimbabwe had the highest estimated under-five diarrheal mortality burden, with 8 deaths and a mortality incidence rate of 0.081 deaths per 100,000 under-five children. Somaliland followed with 7 deaths, while Kenya had the lowest estimated mortality burden with 4 deaths as depicted in Figure 2.

**Figure 2.**
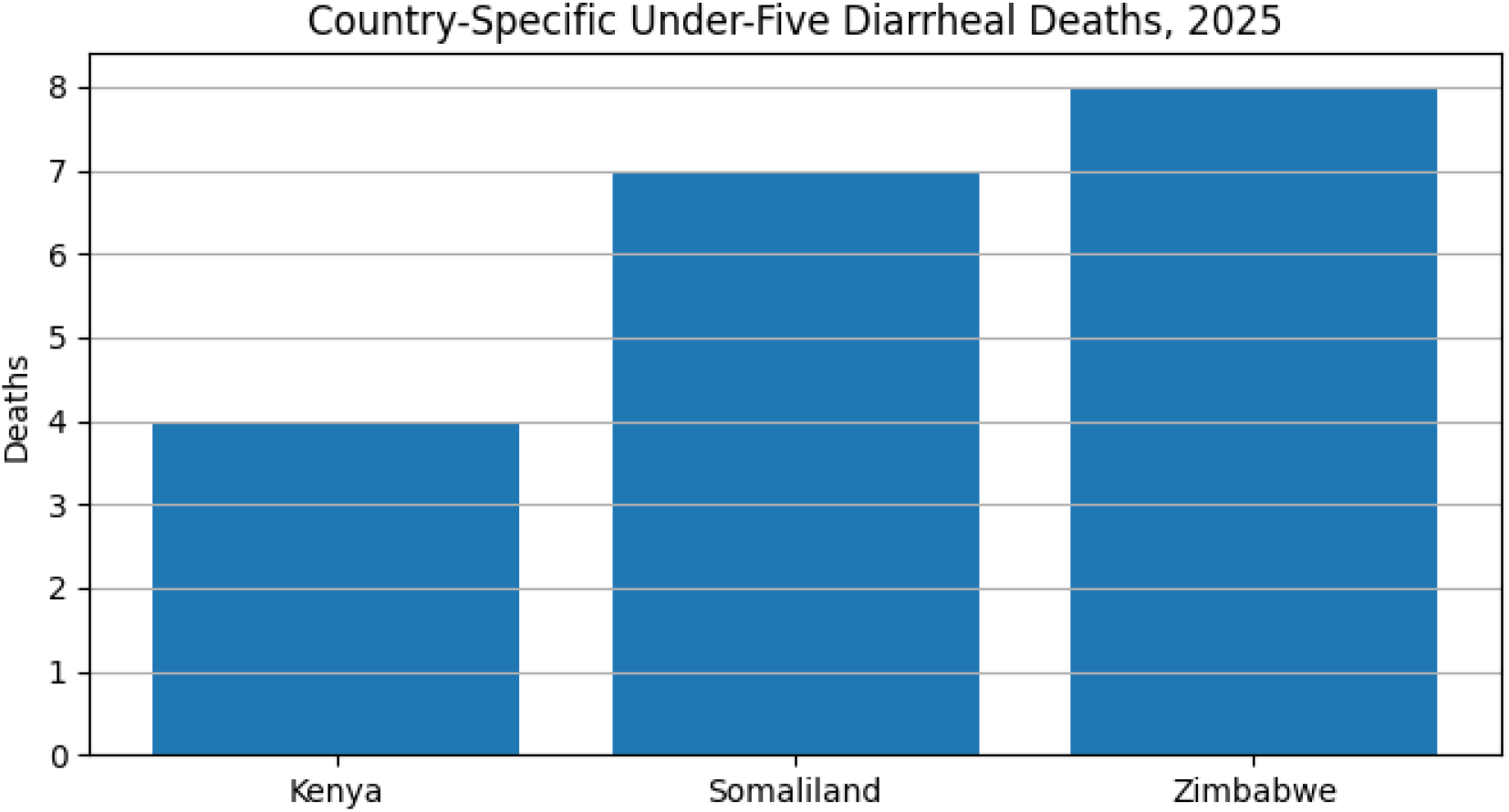
Estimated diarrheal deaths among children under five years of age in Kenya, Zimbabwe, and Somaliland generated using the AFRIDIARRHEA burden estimation framework. Differences in mortality burden reflect variations in disease incidence, healthcare access, environmental risk factors, nutrition status, and underlying population characteristics.

For morbidity, Zimbabwe also had the highest estimated diarrheal case burden, with 553 cases and a morbidity incidence rate of 0.056 cases per 1,000 under-five children as shown in Figure 3. Somaliland was close behind with 542 cases, while Kenya had 356 cases.

**Figure 3.**
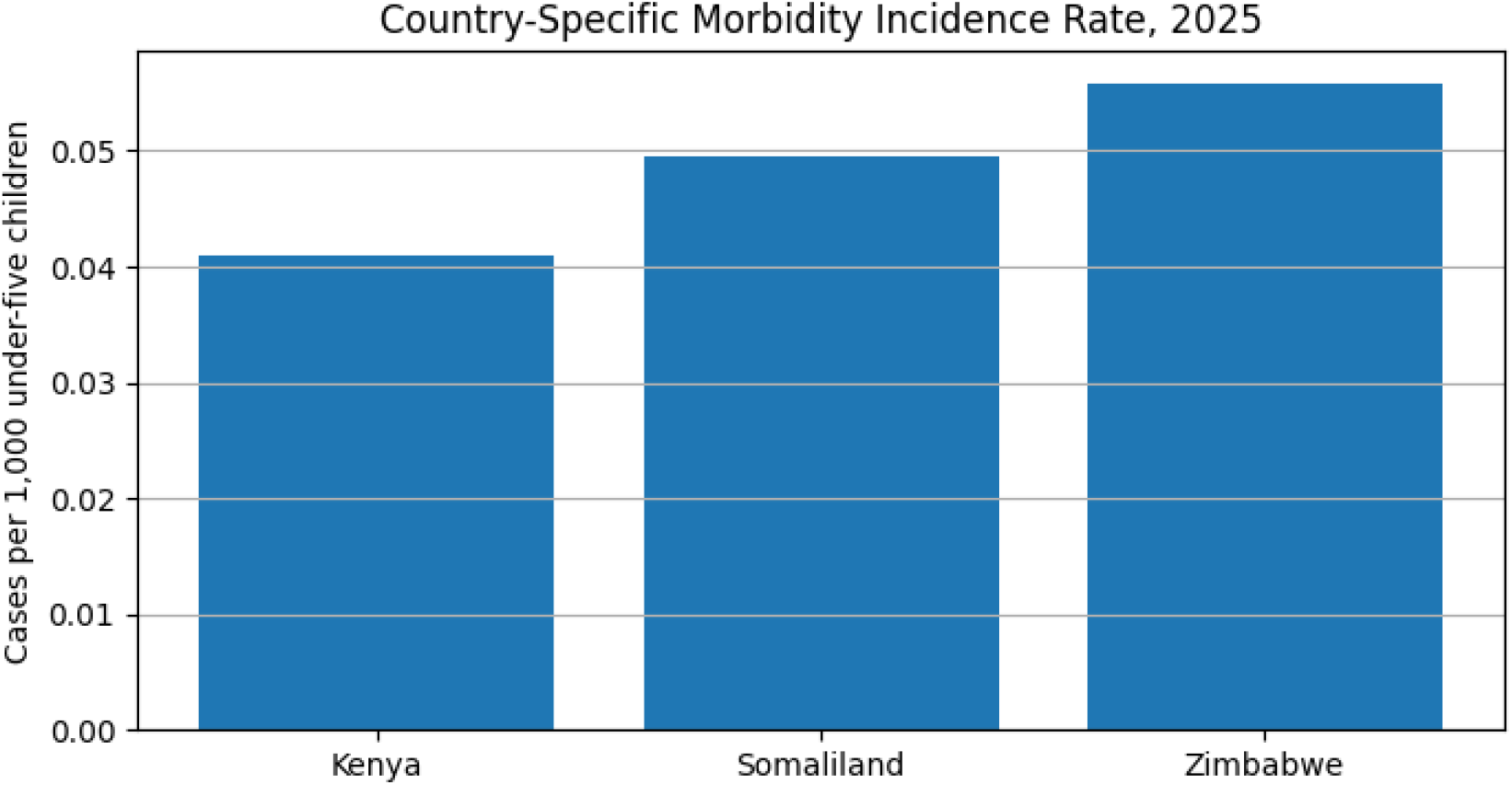
Estimated diarrheal morbidity incidence rates per 1,000 children under five years of age in Kenya, Zimbabwe, and Somaliland. The rates represent the frequency of diarrheal disease occurrence within the under-five population and provide a comparative measure of disease burden across study settings.

Hospitalization burden was highest in Somaliland, with 53 hospitalizations, suggesting that although Zimbabwe had the highest deaths and cases, Somaliland had a greater modeled burden of clinically severe disease requiring hospital care.

### 3.2. Pathogen-Attributed Mortality and Morbidity Burden

Pathogen attribution analysis identified important differences in the contribution of individual enteric pathogens to the overall diarrheal mortality burden among children under five years of age as shown in Figure 4.

**Figure 4.**
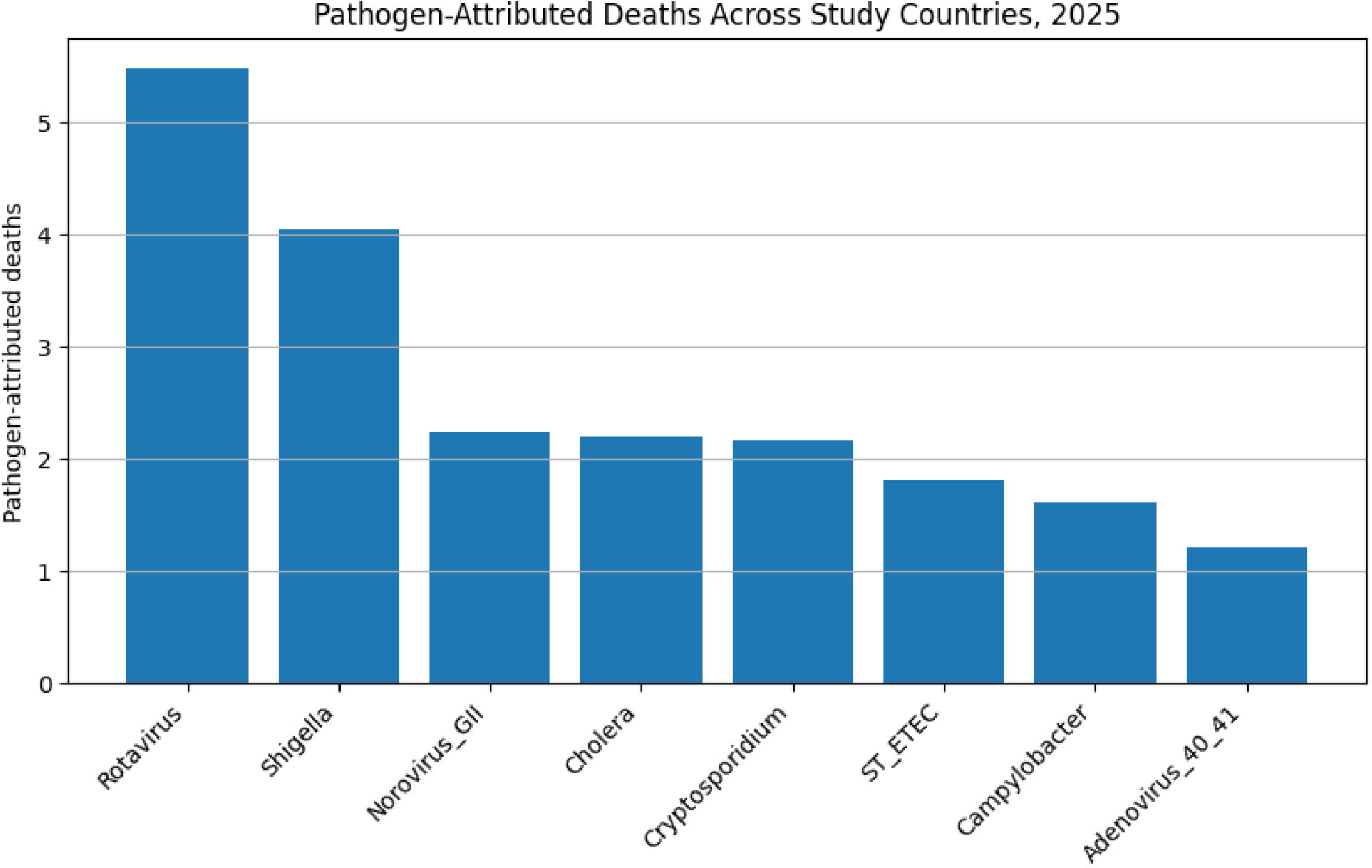
Estimated pathogen-attributed deaths among children under five years of age aggregated across Kenya, Zimbabwe, and Somaliland. Rotavirus accounted for the largest mortality burden, followed by Shigella, while Norovirus GII, Cholera, and Cryptosporidium contributed intermediate mortality burdens. ST-ETEC, Campylobacter, and Adenovirus 40/41 accounted for comparatively smaller proportions of pathogen-attributed deaths.

Aggregated across Kenya, Zimbabwe, and Somaliland, Rotavirus accounted for the highest number of pathogen-attributed deaths, followed by Shigella. Norovirus GII, Cholera, and Cryptosporidium contributed intermediate mortality burdens and exhibited comparable levels of attributable mortality. ST-ETEC and Campylobacter contributed smaller proportions of pathogen-attributed deaths, while Adenovirus 40/41 demonstrated the lowest attributable mortality burden among the pathogens included in the analysis. The pathogen-attributed mortality profile revealed a clear hierarchy of disease burden, with Rotavirus contributing substantially more deaths than any other pathogen. Shigella represented the second most important contributor, whereas Norovirus GII, Cholera, and Cryptosporidium formed a middle-tier group of pathogens associated with moderate mortality burden. The lowest mortality contributions were observed for ST-ETEC, Campylobacter, and Adenovirus 40/41.

These results demonstrate that the burden of childhood diarrheal mortality is distributed across multiple etiologic agents rather than being driven by a single pathogen. Nevertheless, Rotavirus and Shigella collectively accounted for the largest proportion of pathogen-attributed mortality, indicating their dominant role in severe diarrheal disease outcomes within the modeled study settings.

### 3.3. Model Performance

The multimodal fusion framework integrated epidemiological, temporal, geospatial, environmental, satellite-derived, and pathogen-specific predictors to estimate diarrheal disease incidence. Model evaluation demonstrated that the multimodal fusion model achieved the best overall predictive performance, outperforming the Bayesian baseline and individual component models according to RMSE and MAE metrics as shown in Figure 5.

**Figure 5.**
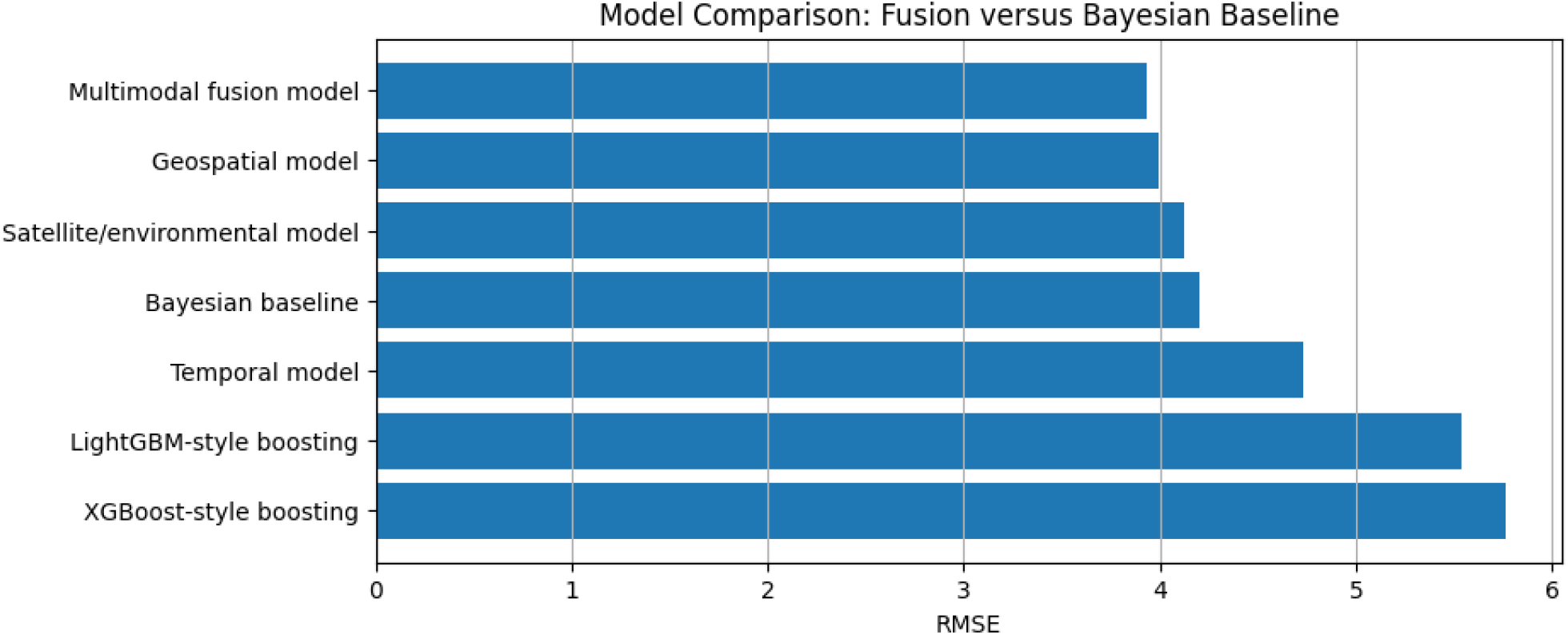
RMSE comparison of the Bayesian baseline, component models, and the multimodal fusion model. Lower RMSE indicates better predictive performance

The fusion architecture combined information from multiple complementary modeling approaches, including boosting models, temporal predictors, geospatial variables, and environmental risk indicators, resulting in improved predictive accuracy and robustness. The fusion weights indicated that the strongest-performing component models contributed most substantially to the final ensemble prediction.

Figure 6 shows the observed versus predicted comparison. It demonstrates that the multimodal fusion model produced predictions that more closely aligned with the line of perfect agreement than the Bayesian baseline.

**Figure 6.**
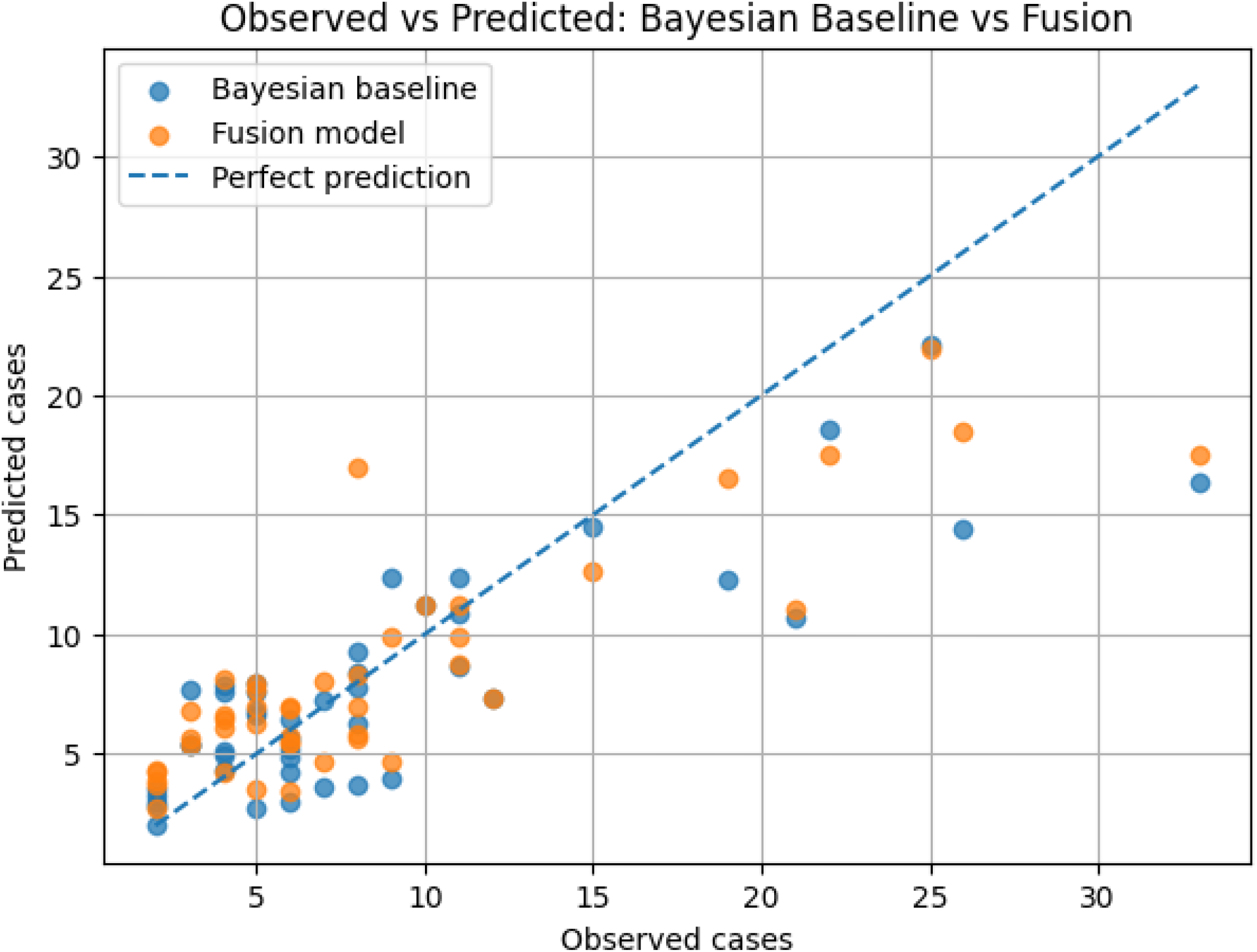
Comparison of observed and predicted case counts. The dashed line represents perfect prediction, with points closer to the line indicating higher predictive accuracy. The multimodal fusion model demonstrated improved agreement with observed cases relative to the Bayesian baseline

### 3.4. Uncertainty, Benchmark Comparison, and Sensitivity Analysis

The AFRIDIARRHEA framework generated point estimates accompanied by 95% uncertainty intervals derived from model prediction error and disagreement among component models within the multimodal fusion architecture as shown in Figure 7.

**Figure 7.**
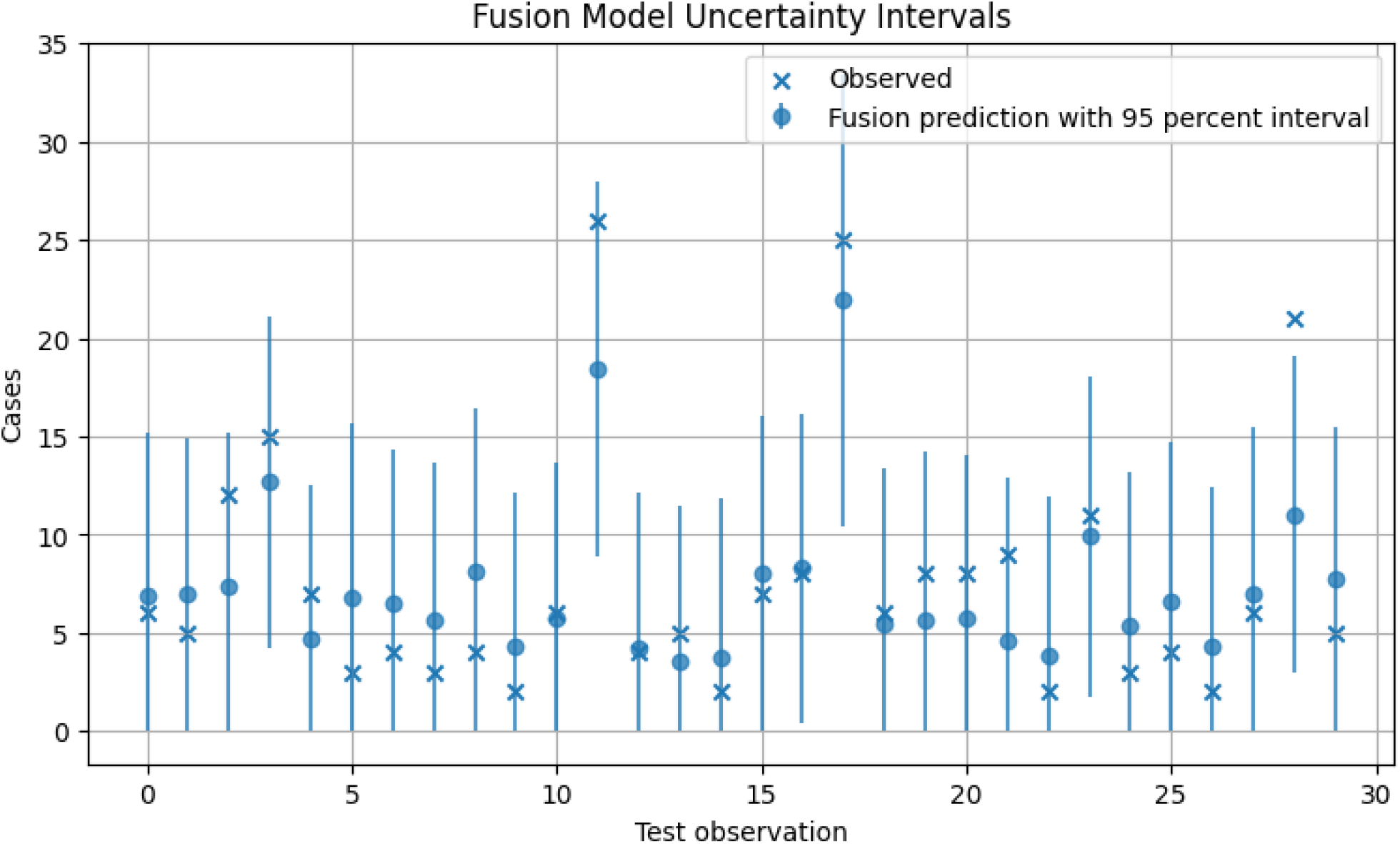
Observed and predicted diarrheal cases with corresponding 95% uncertainty intervals generated by the multimodal fusion model. Error bars represent prediction uncertainty, with wider intervals indicating greater model uncertainty.

The resulting uncertainty intervals demonstrated appropriate empirical coverage of observed diarrheal cases, indicating that the framework effectively quantified predictive uncertainty while maintaining robust forecasting performance. Comparison with external benchmark estimates showed generally close agreement between the AFRIDIARRHEA estimates and reference burden values, with only modest differences observed across countries. Sensitivity analysis further demonstrated that pathogen-attributed mortality estimates responded predictably to variations in pathogen-specific relative risk assumptions, with higher relative risks producing larger attributable fractions and greater estimated mortality burdens.

## 4. Discussion

The country-level burden estimates demonstrated substantial heterogeneity in modeled diarrheal disease outcomes across Kenya, Somaliland, and Zimbabwe. Zimbabwe exhibited the highest mortality and morbidity burden, with the greatest number of diarrheal cases, severe cases, deaths, and incidence rates among children under five, while Somaliland recorded the highest hospitalization burden, indicating a greater proportion of severe cases requiring hospital care. Kenya consistently showed the lowest burden across all indicators. Although these estimates were generated from a synthetic proof-of-concept dataset and should not be interpreted as actual epidemiological values, the findings illustrate the capability of the AFRIDIARRHEA framework to capture country-level differences in disease burden and integrate multiple epidemiological, environmental, healthcare, and demographic risk factors into a unified burden estimation system.

The observed dominance of Rotavirus is consistent with global epidemiological evidence identifying Rotavirus as one of the leading causes of severe childhood diarrhea and diarrheal mortality, particularly among children under five years of age. Although widespread vaccine introduction has reduced disease burden in many countries, Rotavirus remains a major contributor to severe disease outcomes in resource-limited settings where vaccine coverage may be incomplete and healthcare access remains constrained. The substantial contribution of Shigella further highlights its importance as a major etiologic agent associated with severe diarrheal disease and mortality. Shigella infections are often linked to dysentery, dehydration, and malnutrition, factors that increase the risk of adverse outcomes among vulnerable pediatric populations. The elevated mortality burden associated with Shigella reinforces the need for continued surveillance, antimicrobial resistance monitoring, and development of effective preventive interventions. Norovirus GII, Cholera, and Cryptosporidium demonstrated intermediate mortality contributions, suggesting that these pathogens remain important components of the overall diarrheal disease burden. The contribution of Cholera reflects the continued risk posed by waterborne disease transmission, particularly in settings with inadequate water and sanitation infrastructure. Similarly, Cryptosporidium remains an important cause of severe disease among malnourished and immunocompromised children, while Norovirus contributes substantially to community-level transmission and disease incidence. The comparatively lower mortality burden associated with ST-ETEC, Campylobacter, and Adenovirus 40/41 does not diminish their public health significance. These pathogens continue to contribute to morbidity and may serve as important drivers of disease transmission, particularly in specific geographic regions or population subgroups. Their inclusion within the attribution framework provides a more comprehensive assessment of diarrheal disease etiology and supports broader public health planning. Overall, the pathogen attribution findings highlight the multifactorial nature of childhood diarrheal disease and underscore the importance of integrated intervention strategies. The results suggest that vaccination programs targeting Rotavirus, improved WASH infrastructure, enhanced pathogen surveillance, outbreak preparedness measures, and strengthened healthcare access could substantially reduce both morbidity and mortality burden across the study settings.

The superior performance of the multimodal fusion model demonstrates the value of integrating diverse data streams and modeling approaches for estimating diarrheal disease burden in complex infectious disease systems. By combining Bayesian epidemiological structure, nonlinear machine-learning models, temporal forecasting, geospatial analytics, pathogen attribution, and environmental intelligence, the AFRIDIARRHEA framework was able to capture complex interactions among climate variability, pathogen prevalence, healthcare access, sanitation conditions, demographic factors, and spatial heterogeneity more effectively than the Bayesian baseline alone. The findings suggest that multimodal fusion architectures can substantially improve predictive accuracy while preserving epidemiological relevance, highlighting their potential as robust tools for burden estimation, forecasting, and evidence-based public health decision-making in African settings characterized by heterogeneous disease transmission dynamics.

The AFRIDIARRHEA framework generated reliable burden estimates by combining multimodal fusion modeling with uncertainty quantification. Most observed diarrheal case counts fell within the predicted 95% uncertainty intervals, indicating good calibration, while benchmark and sensitivity analyses demonstrated that the estimates were robust to reasonable variations in model assumptions and pathogen-specific risks. These findings highlight the framework’s potential for producing accurate, transparent, and policy-relevant burden estimates for public health decision-making. The observed-versus-predicted analysis demonstrated that the multimodal fusion model achieved closer agreement with observed diarrheal case counts than the Bayesian baseline, indicating improved predictive accuracy across varying levels of disease burden. By integrating epidemiological, temporal, geospatial, environmental, and pathogen-specific information, the fusion framework more effectively captured the complex nonlinear relationships underlying diarrheal disease transmission, resulting in reduced prediction error and more reliable burden estimates than a single-model approach.

The AFRIDIARRHEA framework addresses several limitations of existing diarrheal burden-estimation approaches by integrating epidemiological, environmental, temporal, spatial, and pathogen-specific information within a unified multimodal architecture while explicitly incorporating uncertainty quantification, sensitivity analysis, coinfection handling, and transparent modeling assumptions. These features enable the generation of policy-relevant outputs that can support vaccine prioritization, WASH investment planning, outbreak preparedness, climate-health adaptation, hospital resource allocation, and child survival programs, particularly in LMICs characterized by sparse surveillance systems and incomplete etiologic data.

## 5. Conclusion

This study developed AFRIDIARRHEA, a multimodal fusion framework for estimating childhood diarrheal morbidity, mortality, pathogen-attributed burden, and uncertainty in African settings. By integrating Bayesian epidemiological modeling, machine learning, temporal forecasting, geospatial analytics, environmental intelligence, and pathogen attribution, the framework generated comprehensive burden estimates while capturing complex disease-transmission dynamics. The proof-of-concept application revealed substantial heterogeneity across Kenya, Zimbabwe, and Somaliland, identified Rotavirus and Shigella as major contributors to pathogen-attributed mortality, and demonstrated improved predictive accuracy compared with the Bayesian baseline.

The findings highlight the potential of multimodal fusion architectures to enhance diarrheal burden estimation through the integration of diverse data streams, uncertainty quantification, and transparent modeling assumptions. Although based on synthetic data and simplified modeling components, the framework demonstrates the feasibility of supporting vaccine prioritization, WASH investments, outbreak preparedness, climate-health adaptation, healthcare resource allocation, and child survival programs. Future work should incorporate real-world surveillance, laboratory, mortality, climate, and satellite data, together with full Bayesian inference, to improve accuracy, interpretability, and operational applicability in resource-constrained settings.

## Author Approval Statement

All authors have read and approved the final version of this manuscript and consent to its submission to medRxiv. Each author has made a substantial contribution to the conception, design, data collection, analysis, interpretation, drafting, or revision of the work and agrees to be accountable for all aspects of the study. The authors confirm that the manuscript is original, has not been published elsewhere, and is not under consideration for publication by another journal or preprint server, except as permitted by medRxiv policies. All authors have reviewed the content and approved its public posting on medRxiv.

## Declaration

This study utilized publicly available, anonymized, synthetic, or secondary data sources and did not involve direct interaction with human participants or access to identifiable personal information. Therefore, ethics committee approval and informed consent were not required. Where applicable, all procedures were conducted in accordance with relevant institutional guidelines and regulations.

## Data Availability Statement

The datasets and materials used and/or analyzed during the current study are available from the corresponding author upon reasonable request. Publicly available data sources are appropriately cited within the manuscript.

## References

1. World Health Organization. (2024). Diarrhoeal disease. World Health Organization. https://www.who.int/news-room/fact-sheets/detail/diarrhoeal-disease

2. Kyu, H. H., et al. (2025). Global, regional, and national age-sex-specific burden of diarrhoeal diseases. The Lancet Infectious Diseases.

3. UNICEF. (2024). Diarrhoea remains a leading killer of young children. United Nations Children’s Fund. https://data.unicef.org/topic/child-health/diarrhoeal-disease/

4. GBD 2021 Causes of Death Collaborators. (2024). Global burden of 288 causes of death and life expectancy decomposition in 204 countries and territories and 811 subnational locations, 1990-2021: A systematic analysis for the Global Burden of Disease Study 2021. The Lancet, 403(10440), 2100–2132. 10.1016/S0140-6736(24)00367-2

5. Black, R. E., Perin, J., Yeung, D., Rajeev, T., Miller, J., Elwood, S. E., Platts-Mills, J. A., et al. (2024). Estimated global and regional causes of deaths from diarrhoea in children younger than 5 years during 2000-21: A systematic review and Bayesian multinomial analysis. The Lancet Global Health, 12(6), e919–e928. 10.1016/S2214-109X(24)00078-0

6. Kotloff, K. L., Nataro, J. P., Blackwelder, W. C., Nasrin, D., Farag, T. H., Panchalingam, S., Wu, Y., Sow, S. O., Sur, D., Breiman, R. F., Faruque, A. S. G., Zaidi, A. K. M., Saha, D., Alonso, P. L., Tamboura, B., Sanogo, D., Onwuchekwa, U., Manna, B., Ramamurthy, T., … Levine, M. M. (2013). Burden and aetiology of diarrhoeal disease in infants and young children in developing countries (The Global Enteric Multicenter Study, GEMS): A prospective, case-control study. The Lancet, 382(9888), 209–222. 10.1016/S0140-6736(13)60844-2

7. Platts-Mills, J. A., Babji, S., Bodhidatta, L., Gratz, J., Haque, R., Havt, A., McCormick, B. J. J., McGrath, M., Olortegui, M. P., Samie, A., Shakoor, S., Mondal, D., Lima, I. F. N., Hariraju, D., Rayamajhi, B. B., Qureshi, S., Kabir, F., Yori, P. P., Moulton, L. H., … Houpt, E. R. (2015). Pathogen-specific burdens of community diarrhoea in developing countries: A multisite birth cohort study (MAL-ED). The Lancet Global Health, 3(9), e564–e575. 10.1016/S2214-109X(15)00151-5

8. Troeger, C., Blacker, B. F., Khalil, I. A., Rao, P. C., Cao, J., Zimsen, S. R. M., Albertson, S. B., Deshpande, A., Farag, T., Abebe, Z., Adetifa, I. M. O., & GBD 2017 Diarrhoeal Disease Collaborators. (2020). Estimates of the global, regional, and national morbidity, mortality, and aetiologies of diarrhoeal diseases: An analysis of the Global Burden of Disease Study 2017. The Lancet Infectious Diseases, 20(8), 909–948. 10.1016/S1473-3099(19)30401-3

9. Carlton, E. J., Eisenberg, J. N. S., Goldstick, J., Cevallos, W., Trostle, J., & Levy, K. (2016). Heavy rainfall events and diarrhea incidence: The role of social and environmental factors. American Journal of Epidemiology, 179(3), 344–352. 10.1093/aje/kwt279

10. Chen, T., & Guestrin, C. (2016). XGBoost: A scalable tree boosting system. In Proceedings of the 22nd ACM SIGKDD International Conference on Knowledge Discovery and Data Mining (pp. 785–794). Association for Computing Machinery. 10.1145/2939672.2939785

11. Ke, G., Meng, Q., Finley, T., Wang, T., Chen, W., Ma, W., Ye, Q., & Liu, T.-Y. (2017). LightGBM: A highly efficient gradient boosting decision tree. In I. Guyon et al. (Eds.), Advances in Neural Information Processing Systems (Vol. 30, pp. 3146–3154). Curran Associates, Inc.

